# A Retrospective Case Study of Successful Translational Research: Cardiovascular Disease Risk Assessment, Experiences in Community Engagement

**DOI:** 10.1101/2023.09.26.23296161

**Authors:** Michael E. Bales, Jifeng Zhu, Christine A. Ganzer, Farid Aboharb, Allegra Keeler, Krista A. Ryon, Brett J. Ehrmann, Julianne Imperato-McGinley, the H2H Consortium

## Abstract

In underserved communities in New York City, uninsured adults encounter a greater risk of cardiovascular disease and diabetes. The Heart-to-Heart Community Outreach Program (H2H) is addressing these disparities by providing screenings for diabetes and other cardiovascular disease risk factors, fostering community engagement in translational research at our CTSC. Screening events are hosted in partnership with community faith-based institutions. Participants provide medical history, complete a survey, and receive individualized counseling by clinicians with referrals for follow-up care. The population served is disproportionately non-white, uninsured, with low-income, and underserved. The program empowers participants to make beneficial lifestyle changes using myriad strategies to reach those most in need. This required strong foundational program leadership, effective inter-institutional collaboration, and maintaining of community trust. Leveraging partnerships with faith-based institutions and community centers in at-risk NYC neighborhoods, H2H addresses the increasing burden of diabetes and cardiovascular disease risk factors in vulnerable individuals and provides a model for similar initiatives.

## Introduction

The Weill Cornell Clinical and Translational Science Center (CTSC)[1,2], with two of its partners – NewYork-Presbyterian Hospital (NYP)[3], and Hunter-Bellevue School of Nursing (HBSON)[4] – initiated the Heart-to-Heart Community Outreach Program (H2H) in 2010 to help address healthcare disparities contributing to cardiovascular disease in underserved communities of New York City (NYC). A free program, H2H brings the clinic to the community by providing health screenings in NYC’s underserved and minority communities. Sites are open to the public and hosted by the CTSC Community Outreach Core’s network of faith-based churches and senior centers[5].

The overarching goal is to help participants understand their current health status and provide early intervention through on-site consultations and connections to resources for follow-up healthcare. The primary aims include identifying those with diabetes, hypertension, obesity, and hyperlipidemia, and determining prevalence of these conditions among the underserved in NYC. The program enables significant community engagement in the translational research process at our CTSC.

Along with medical initiatives, the program includes educational training for the next generation of healthcare professionals to work effectively in multidisciplinary, patient-centered teams in underserved areas[6]. The multifaceted team consists of undergraduate students working alongside medical students, nursing students, physician assistants, and dietitian students under faculty supervision.

Participants are assessed for height, weight, body mass index (BMI), blood pressure, and waist circumference, as well as biochemical measurements (blood glucose, hemoglobin A1c, and a complete lipid panel – HDL, LDL, total cholesterol, and triglycerides). A clinician counsels the participant, explains the results, and provides individualized follow-up information. Dieticians are frequently available to provide counseling when needed. Additionally, socioeconomic and demographic history questionnaires are completed to understand factors contributing to an increased disease burden in these populations. By engaging local communities, H2H is understanding how behavior, knowledge of cardiovascular disease (CVD), and systemic factors impact differences in risk and future healthcare decision-making.

The objectives of this translational science case study are to 1) quantify the health status of those participating in H2H screenings, 2) identify the socioeconomic, health access, and health-related barriers disproportionately promoting the onset of CVD and diabetes; and 3) describe how the program has evolved in response to its findings throughout its first 12 years, and plans for the future.

## Background

With 600,000 uninsured residents[7], NYC has the largest uninsured urban population in the United States. Uninsured residents encounter a greater risk of developing CVD and diabetes[8]. Over the past decade, CVD was the leading cause of death in NYC, and diabetes has often ranked in the top five[9]. This is partially the result of multiple barriers facing underserved communities – social, cultural, and economic health determinants resulting in poor access to preventative healthcare. Furthermore, the COVID-19 pandemic disproportionately affected vulnerable communities and exacerbated systemic inequities in healthcare[10].

In 2018, healthcare spending and lost productivity due to CVD exceeded $400B[11], making it the costliest disease in the US. Despite decades of steady decreases in overall CVD risk, racial, geographic, and socioeconomic health disparities persist among specific subgroups. Screening for modifiable risk factors, such as high blood pressure, obesity, diabetes, and elevated cholesterol, is critical for CVD risk reduction, especially through early intervention[12]. However, individuals in low-income and medically underserved communities often encounter barriers in the healthcare system[13]. Given NYC’s large uninsured, underinsured, and underserved population, innovative community-academic partnerships help address these barriers and promote health equity[14]. Prevention and early intervention via lifestyle modification and/or first-line medications are successful and cost-effective – preventing an additional 3.2 cases of diabetes per 100 person-years in at-risk populations[15]. Similarly, the American Heart Association cites numerous exercise programs and found that these interventions effectively reduced systolic and diastolic blood pressure, increased HDL, decreased LDL and triglycerides, and reduced insulin resistance and glucose intolerance[16]. Consequently, relatively simple interventions, performed consistently and with appropriate follow-up, could prevent significant amounts of disease and disability in underserved populations.

The CTSC H2H program was developed with input from the Weill Cornell CTSC’s Community Advisory Board (CAB) and community partners. The H2H executive board of students and faculty coordinates research activities and medical protocol decisions, and community leaders promote and host events, determining the date/time of events based on community needs. Over the years, additional services such as nutrition counseling and influenza vaccinations were added at the request of community partners, and several leaders from partner sites became members of the CAB.

### Chronology of developmental milestones and process markers

#### The early years: identifying the target community and establishing the community partnership

H2H was initiated by two MD/PhD students at WCM to combat CVD in underserved NYC populations[17]. Inspired by their experience at Weill Cornell Community Clinic, they wanted to “bring the clinic to the community” and approached the PI of the CTSC. With help from CTSC External Advisory Board member Reverend Patrick O’Connor, a pilot event was organized at his church, which serves predominantly low-income African American and Hispanic neighborhoods.

The program gained momentum when it caught the attention of a State Senator’s chief of staff at the first event; additional events were scheduled in collaboration with the senator’s office. Simultaneously, CTSC staff worked with the CTSC CAB to extend the program’s reach.

Ultimately, word of mouth through faith-based networks proved the most effective method for establishing new partnerships. Successful early events led to an abundance of requests from other faith-based organizations in the third year. Noting the impact and collaborative process of H2H screening events, the number of requests for events steadily grew to, and remains, dozens per year.

#### Roles of key stakeholders in the program’s growth and development

With the support of community partners, including clergy members, H2H gradually transitioned from its loosely defined goal of reaching out to the underserved to holding screening events in low-income minority groups in NYC. Community leaders were essential in advocating for neighborhood events and determining screenings offered. Over time, the program continued to gain attention and recognition, and just three years after H2H was initiated, the number of requests for events began to exceed the program’s capacity^1^.

#### Building community trust as an annual healthcare tradition

At least two unanticipated community-level effects have emerged from H2H. First, despite emphasis by program organizers to the contrary, some participants have expressed that the program serves as their primary source of healthcare and is regarded by some community members as an annual tradition. While H2H was never intended for this purpose, some communities have large numbers of undocumented immigrants who have difficulty obtaining healthcare and health insurance.

A second effect of the program is development of trust between the community and H2H representatives, and the community and the CTSC. H2H volunteers have perceived an increase in trust among community members after multiple consecutive annual events at a site, and these annual events have likely helped assuage concerns of participants that they are being taken advantage of by the medical establishment (a perception that has been well-documented in research involving underserved communities[18,19]).

Establishing and maintaining trust has led to increased participation among community members with other CTSC-related research initiatives. These events have also served as an infrastructure to aid CTSC investigators in recruiting healthy volunteers for other studies^2^. Overall, multiple CTSC-supported research studies have successfully recruited large numbers of underrepresented minority participants – a traditionally difficult population to recruit. In addition, some community members have accepted invitations to serve on the CTSC’s CAB, where they assist in reviewing grant applications and participate in CTSC grant funding decisions. This way, H2H serves as a bridge, allowing community engagement throughout the translational research process.

### Factors that were essential to the collaboration (facilitators)

Facilitators that have been essential to the collaboration include effective inter-institutional leadership and participation and building and maintaining community trust.

A core team administers the H2H program through the WCM CTSC’s Community Engagement and Research Component. The Managing Director of the component (J.Z.) has been involved since the program’s inception, serving as a consistent community point of contact and knowledge over the years. A specific aim of the program is to introduce students to the concept of multidisciplinary team-based health care.

Volunteers are recruited from WCM, NewYork-Presbyterian Hospital, and Hunter College (HC). WCM volunteers include medical students, physician assistants, and dietician students. HC volunteers include nursing students and undergraduates from the college interested in exploring the health professions. In addition, the program recruits licensed medical professionals (physicians, nurse practitioners, and physician assistants) from WCM and HC to carry out educational counseling based on current guidelines[20–22].

For the first few years, the role of the CTSC was to provide funding as well as regulatory, administrative, and logistical support. With the growth of the program, the CTSC expanded to handle most of the program while also helping lay out scientific goals in detail.

Community leaders embraced the H2H program because it was the first comprehensive program most had encountered. While some programs offered singular point-of-care testing (lipid, blood pressure, mammograms, etc.), none were integrated into a comprehensive program that provided multiple tests, included referrals to follow-up care, insurance enrollment, and, most importantly, medical consultations on-site. The annual return of the program further cemented this trust^3^.

### Challenges and barriers to implementation

There were several key challenges encountered when developing the H2H Program, including institutional hurdles prior to launching, recruitment of attending physicians, ongoing funding, and transitions in leadership.

#### Institutional and IRB-related challenges

The first challenge was that organizers planned to involve students in our CTSA consortium. Organizers reasoned that the number of students would be large and frequently changing, which could burden both the project and the IRB with paperwork. The IRB eventually agreed that any healthcare students whose role was limited to volunteering at medical screening events would not need to be listed on the IRB protocol, but requested a record of those involved.

The second main institutional hurdle was balancing the program’s community service goals with the strict requirements of a public health research initiative, resulting in two challenges – one pertaining to the consent of participants and the other to institutional risk and insurance.

For the program’s first five years, participants were recruited anonymously to help mitigate the complexities of the recruitment process. By keeping participants anonymous, the IRB initially determined that verbal consent would be sufficient, allowing the program to obtain consent from a large number of participants (over 100 in some cases) in a very short period (on the order of an hour or two). However, this precluded the possibility of following up with participants to determine outcomes, severely hampering research efforts. We recently resolved this issue by working with the IRB to classify the program’s survey as a QA/QI project.

An additional challenge arose from medical care being provided off-campus at locations unaffiliated with the organizers’ medical college or hospital network. This was the first time any program had approached the institution to conduct medical activities outside the institution, and as a result, liability and insurance coverage had not been established. In a series of meetings involving H2H Campaign leadership, the leadership of the IRB, and the risk management office, it was ultimately determined that if an attending physician affiliated with the hospital network were present on-site at all events to oversee medical activities, their insurance would cover medical activities and student volunteers.

Additionally, the IRB and Risk Management required that communications with participants would be limited to identifying likely (but not confirmed) diagnoses and that participants would be referred to follow-up care for confirmation. Overall, during program development, obtaining authorization from relevant institutional offices required substantial effort.

#### Recruitment of attending physicians

The program has been fortunate to have support of several highly dedicated medical faculty volunteers devoting their time and effort. A key strategy employed by program leadership has been to obtain a list of new attending physicians annually and to reach out to them by email. Organizers have found that newly hired attending physicians building their practices may have more interest to volunteer on weekends. The program has received strong support from the Chair of the Primary Care Department at WCM, with most medical directors and volunteer physicians recruited from this group. Another successful strategy has been through medical students who identify prospective faculty volunteers through their coursework.

#### Ongoing funding

The program was initially funded through a two-year pilot award from the CTSC, then via a grant from the AstraZeneca Foundation for two years. However, for most of its existence, funding has been through the community engagement and research component of the CTSC^4^.

#### Transitions of leadership and program continuity

As students eventually graduate, transfer schools, or leave for other reasons, transition of leadership and program continuity is often a concern in student-run projects. While H2H founders were MD/PhD students who were present for many years, organizers realized early on that continuity was needed. To meet these challenges^5^, the Managing Director of the CTSC community component (J.Z.) has been involved in this project since its inception, taking charge and serving as a consistent organizer and community contact and aggregating institutional knowledge over the years.

#### Additional challenges

Two significant changes were made to the program after the first event. First, recruiting of Spanish-speaking staff and volunteers to address the large non-English speaking Hispanic population in NYC. Second, the addition of undergraduate student volunteers to escort participants through the screening process and administer study surveys to prevent participant confusion and ensure accurate recordkeeping.

There were also two activities that organizers anticipated would be challenging for the long term, but did not turn out to be: identifying community partners and recruiting student volunteers. As mentioned earlier, screening requests were abundant by the third and fourth years, aided by a positive relationship with and annual returns to screening sites. As for recruitment of student volunteers, organizers initially anticipated challenges, as events require dozens of student volunteers. Although it is more difficult to find student volunteers at specific times of the year (the month of graduation and the summer, in particular), organizers have never experienced a shortage^6^.

#### Limitations of the study

The major limitation of the program is that the demographic and health-related data collected are based on a convenience sample, a significant barrier to the application of rigorous experimental models. A second limitation is that the program has not yet incorporated protocols to track the progress of individual participants over time. The program is currently planning an expansion to implement this.

### Screening protocol

The WCM Institutional Review Board approved the H2H study for generally healthy individuals over 18. WCM team members obtain study consent and pair participants with a student navigator who escorts them through the screening process. Participants provide self-reported demographics, medical history, and complete a survey. A licensed clinician counsels participants on individualized CVD risk factors in conjunction with trainees in medicine and nutrition, provides educational materials on appropriate lifestyle and behavioral modification, and resources for follow-up care. Participants who are uninsured or do not have access to primary care services are given a list of free or sliding scale-based federally-qualified community health clinics or are referred to the Weill Cornell student-run free medical clinic. Individuals who cannot provide informed consent or refuse to participate in the research study still receive all screening and counseling services.

Blood samples are taken from a single finger stick to perform a lipid profile^7^, hemoglobin A1c^8^, and blood glucose. Blood pressure, height, weight, and waist circumference are measured, and body mass index (BMI) is calculated. The student navigator records the participant’s data on an informational handout given to the participant, as well as in a tablet that connects to the study’s research database.

Demographic, anthropomorphic, and health screening data were entered into a database [23], aggregated, and reported as summary statistics (counts, percentages, means, and standard deviations). Comparisons were made using corresponding publicly available data from NHANES. Thematic maps are created by overlaying participants’ home ZIP codes as a density function atop a NYC public-domain map (see Figure 1, below).

### Current status – implementation and dissemination

From 2010 to 2020, the program held 130 events and performed 5,959 screenings at sites across NYC. Most sites have become partners, some as long as 12 years. Activities were temporarily suspended in 2020 due to the pandemic. Beginning in mid-2021, an abridged version of the screenings that minimizes contact was offered at CTSC-supported COVID-19 vaccine sites. In mid-2022 the program resumed normal screenings with reduced capacity. We fully restored all program activities beginning in 2023.

**Table 1.** Demographics and healthcare utilization patterns of participants in the Heart-to-Heart Community Outreach Program. While the total number of unique participants was 4,300, the number of participants (n) for each variable changes due to repeat screenings and changes in questionnaire content at H2H screenings over time.

| *Item* | *Count* | *Percent (%)* |
| --- | --- | --- |
| ***Number of Events*** | *130* |  |
| ***Number of Screenings*** | *5959* |  |
| ***Sex (n=5834)*** |  |  |
| *Male* | *2148* | *36.8* |
| *Female* | *3682* | *63.1* |
| *Other* | *1* | *0.017* |
| *Prefer not to state* | *3* | *0.051* |
| ***Age (n=5774)*** |  |  |
|  | ***Mean*** | ***Standard Dev*** |
|  | 54.3 years | 39.6 |
| **Race (n=5613)** |  |  |
| Black | 3204 | 57.0 |
| Latino | 773 | 13.8 |
| Asian | 863 | 15.4 |
| White | 456 | 8.12 |
| American Indian/Alaskan Native | 44 | 0.8 |
| Pacific Islander | 19 | 0.3 |
| Prefer not to state | 72 | 1.3 |
| Other | 182 | 3.2 |
| **Ethnicity (n=5593)** |  |  |
| Hispanic | 995 | 17.8 |
| Non-Hispanic | 4598 | 82.2 |
| **Income (n=4264)** |  |  |
| \$0-\$20,000 | 1378 | 32.3 |
| \$20,001-\$50,000 | 1120 | 26.3 |
| Over \$50,001 | 1054 | 24.7 |
| Decline to answer | 712 | 16.7 |
| **Insurance status (n=5959)** |  |  |
| None | 1698 | 28.5 |
| Medicaid | 194 | 3.26 |
| Medicare | 346 | 5.81 |
| Private | 501 | 8.41 |
| Other | 154 | 2.58 |
| Yes, non-specific<sup>9</sup> | 3066 | 51.5 |
| **Diabetes Mellitus (n=5842)** |  |  |
| At presentation | 277 | 4.74 |

<sup>9</sup> For the first six years of the H2H program, participants were not asked to specify type of insurance.

|  |  |  |
| --- | --- | --- |
| *Previous diagnosis* | 917 | 15.7 |
| *If diagnosed, sub-optimal control? (Includes those on and off medication)<sup>10</sup>* | 380 | 41.4 |
| *If diagnosed, on medication?* | 695 | 75.8 |
| **Dyslipidemia (n=5699)** |  |  |
| *At presentation* | 1651 | 29.0 |
| *Previous diagnosis* | 2149 | 37.7 |
| *If diagnosed, sub-optimal control? (Includes those on and off medication)<sup>11</sup>* | 1335 | 62.1 |
| *If diagnosed, on medication?* | 1018 | 47.4 |
| **Hypertension (n=5728)** |  |  |
| *At presentation* | 1766 | 30.8 |
| *Previous diagnosis* | 2330 | 40.7 |
| *If diagnosed, sub-optimal control? (Includes those on and off medication)<sup>12</sup>* | 1819 | 78.1 |
| *If diagnosed, on medication?* | 1737 | 74.5 |
| **Body Mass Index (n=5829)** |  |  |
| *Obese (BMI $\geq 30$)* | 1985 | 34.1 |
| *Overweight (BMI 25-29.9)* | 2121 | 36.4 |
| *Normal Weight (BMI 18.5-24.9)* | 1655 | 28.4 |
| *Underweight (BMI <18.5)* | 68 | 1.17 |
| **ER usage in last 6 mo. (n=1577)** |  |  |
| *None* | 1342 | 85.1 |
| *1-2* | 208 | 13.2 |
| *3-5* | 21 | 1.33 |
| *>6* | 6 | 0.38 |

<sup>10</sup> Incident cases (and suboptimal control of) diabetes: HbA1c >7.
<sup>11</sup> Incident cases (and suboptimal control of) dyslipidemia: total cholesterol >200, LDL>130, HDL<40, or triglycerides>150.
<sup>12</sup> Incident cases (and suboptimal control of) hypertension: BP >130/80.

Healthcare utilization
|  |  |  |
| --- | --- | --- |
| Have not seen a doctor in the last year (n=5761) | 1052 | 18.3 |
| Avoided seeking medical care in the last year (n=3260) | 2003 | 61.4 |

#### Health status of the disadvantaged communities participating in H2H screenings

The mean age of participants screened was 54.3 years (SD 39.6). 63.1% were female, 91.9% of participants were non-white, and 28.5% were uninsured. 18.3% had not seen a doctor in the past year. Additionally, 61.4% of those screened reported not seeking necessary care for a medical problem in the past year. Annual income revealed that 32.3% reported less than $20k, 26.3% between $20-$50k, 24.7% above $50k. 16.7% declined to answer.

**Figure 1.**
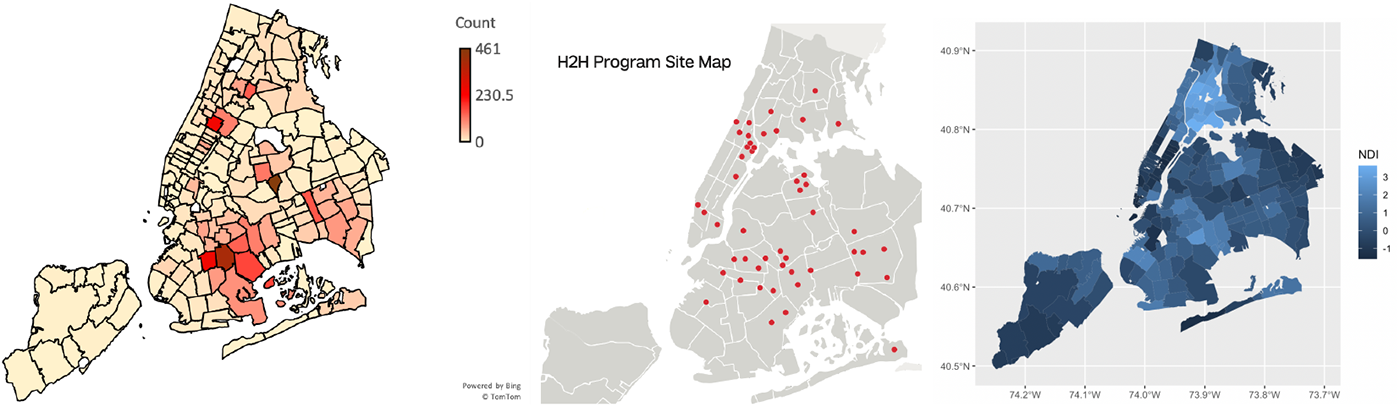
1A. A thematic map showing the geographical distribution of H2H participants throughout the five boroughs of NYC. 1B. A map of sites participating in the Heart to Heart Community Outreach Program (see acknowledgments for a list of sites). 1C. A map of the Neighborhood Deprivation Index for NYC (light blue indicates underserved neighborhoods).

Disease prevalence revealed the following: 40.7% of participants were previously diagnosed with hypertension, with 78.1% having suboptimal control (BP >130/80), despite the fact that 74.5% were on medication. 15.7% of the participants were previously diagnosed with diabetes, with 41.4% having suboptimal control (HbA1c >7), despite the fact that 75.8% of participants with known diabetes were on medication. 37.7% were previously diagnosed with dyslipidemia, and 62.1% had suboptimal control (total cholesterol >200, LDL>130, HDL<40, or triglycerides>150), even though 47.4% were on medication. 34.1% of participants were obese (BMI > 30).

Figure 2 compares the prevalence of selected conditions based on statistics from the NYC Department of Health and Mental Hygiene to that of H2H Program participants.

For those without a previous diagnosis, 30.8% had hypertensive blood pressure, 4.7% had elevated blood sugar (HbA1c >6.5), and 29.0% had dyslipidemia at the time of screening.

**Figure 2.**
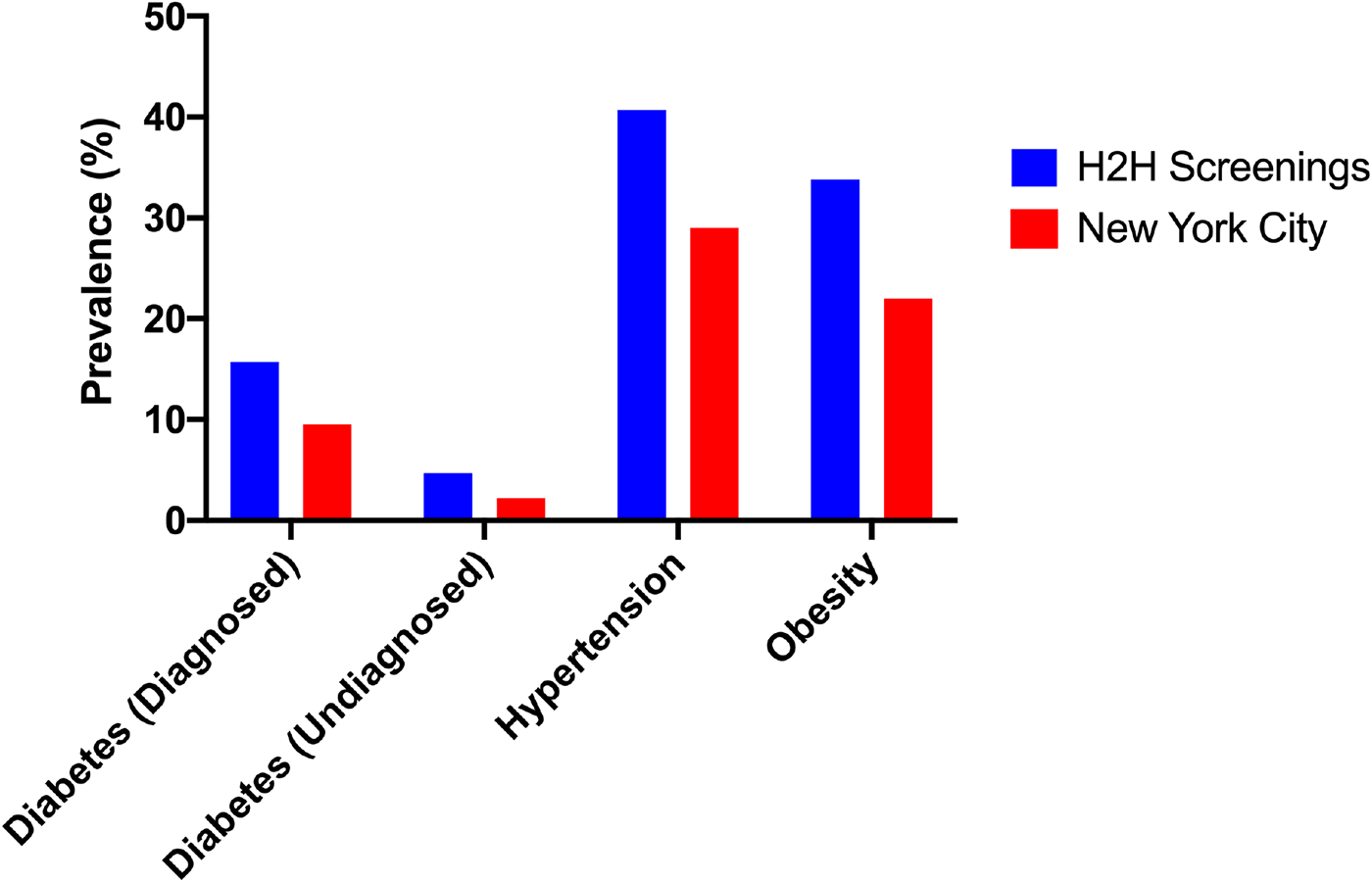
Bar chart comparing the prevalence of these conditions based on statistics from the New York City Department of Health and Mental Hygiene for adults in New York City as a whole to that of H2H Program participants. Undiagnosed diabetes refers to patients who were not aware or did not report having diabetes but who nonetheless met laboratory criteria for diagnosis.

### Further developments

Looking ahead, program leadership has recognized that examining patient-centered health outcomes via long-term follow-up will be a pivotal next step to understand and refine how H2H screenings can become even more impactful to participants in the future.

#### Sharing the results of the research with the community

Medical students have presented the H2H Program at several conferences and events. The CTSC provides updates regularly to its External Advisory Board, as well as its CAB. Updates on the program have also been shared nationally at meetings of sub-committees of the National Center for Advancing Translational Sciences (NCATS) [24].

#### Implications for clinical practice, health education, public health policy, and future research

Based on their experiences interacting with the community over the past 12 years, program organizers have noted significant barriers to receipt of high-quality medical care within underserved communities served by the program. Many participants lack health insurance – a well-recognized problem in underserved communities. However, even among those insured, many perceived their routine interactions with primary care providers as suboptimal. Many, for example, noted that encounters with volunteer providers at the H2H events (lasting approximately 15 minutes, but in more complex cases exceeding 30 minutes) were significantly longer than those at their primary care provider. The ability to speak extensively with an attending physician or nursing faculty member was a new experience for most, if not all, participants.

From an institutional perspective, establishing a large and long-term community-based participatory research network has required long-term commitment. Benefits of the program have extended to the host institution’s research infrastructure. By providing free health screenings, free health education, and other services to these communities, organizers have developed trust and commitment from these communities, enabling them to contribute to the translational research process at the CTSC. They have become members of the CAB, partners in research studies with CTSC principal investigators, and applied for CTSC Community Grants. Through the community network H2H helped develop, many CTSC-supported investigators have had the opportunity to involve underserved communities in their research.

## Conclusion

The Heart-to-Heart Community Outreach Program focuses on “bringing the clinic to the community.” By leveraging strong partnerships with faith-based institutions and community centers in at-risk NYC neighborhoods, the program breaks down the barriers to engaging with the medical establishment and helps to detect and address the increasing burden of diabetes and CVD risk factors in the most vulnerable individuals.

Populations served by the program have been disproportionately non-white, uninsured, low-income, and underserved within the healthcare system. The burden of previously known CVD risk factors is high, and testing has revealed that many of these conditions have been newly discovered or poorly controlled from a medical standpoint. By fostering multidisciplinary and cross-institutional academic-community partnerships, the program has empowered individuals with a more detailed knowledge of their health status, facilitated positive lifestyle modifications, and provided access to medical care, further addressing health risk factors.

By characterizing the community served by the initiative and describing how the program has developed over its 12-year history, we are now preparing a plan for the future. This includes long-term tracking of participants and other measures so that we can continue to optimize this critical community-based program. We hope our experience will provide useful insights to those involved in similar initiatives.

## Data Availability

All data produced in the present study are available upon reasonable request to the authors

## Acknowledgements

We wish to acknowledge the following community and faith-based organizations for their participation in the H2H Program:

Abyssinian Baptist Church, 132 Odell Clark Place, New York, NY

Agape Love Christian Center, 1023 Allerton Ave., Bronx, NY

Alive Ministry, 1050 Beach 21st, Far Rockaway, NY

Atlantic Center Mall, 625 Atlantic Ave. #B3, Brooklyn, NY

Boys & Girls Club, 550 Balcom Ave., Bronx, NY

Caldwell Temple African Methodist Episcopal Zion Church, 1288 Rev. James A. Polite Ave., Bronx, NY

Child Development Support Corp, 802 Kent Ave. #804, Brooklyn, NY

Chinese-American Planning Council, 4812 9th Ave., Brooklyn, NY

Christian Fellowship, 777-779 Schenectady Ave., Brooklyn, NY

Claremont Neighborhood Center, 489 East 169th St., Bronx, NY

Community Health & Resource Fair, 115th St. between 1st and 2nd Avenues, New York, NY

Community Health & Resource Fair, 679 Riverside Drive, New York, NY

Day of Hope, 115th St. between 1st and 2nd Avenues, New York, NY

Dunk the Junk, Beacon High School in Harlem, 1820 Lexington Ave., New York, NY

Emmanuel Baptist Church, 279 Lafayette Ave., Brooklyn, NY

Ephesus 7th Day Adventist Church, 101 W 123rd St., New York, NY

First Baptist Church, 100-10 Astoria Blvd., East Elmhurst, NY

First Presbyterian Church in Jamaica, 89-60 164th St., Jamaica, NY

Flatbush Reformed Church, 890 Flatbush Ave., Brooklyn, NY

Frederick Douglas Academy I, 2581 Adam Clayton Powell Jr., New York, NY

God’s Battalion of Prayer Church, 661 Linden Blvd., Brooklyn, NY

Grace Baptist Church, 223 New Jersey Ave., Brooklyn, NY

Grace United Methodist Church, 33 Saint Johns Pl., Brooklyn, NY

Hanson Place SDA Church, 88 Hanson Pl., Brooklyn, NY

Hebron SDA Church, 1256 Dean St., Brooklyn, NY

Immaculate Conception Church of the Blessed Virgin Mary, 601 Melrose Ave., Bronx, NY

IPRHE/Bronx River Senior Center, 1619 E. 174th St., Bronx, NY

Ismaili Center, 92-68 Queens Blvd., Rego Park, NY

La Familia Adult Day Center, 3014 Fulton St., Brooklyn, NY

Leviticus Church of God, 114-12 Van Wyck Expy., Queens, NY

Linden SD-Adventist Church, 2820 137th Ave., Springfield Gardens, NY

Mamre Seventh Day Adventist Church, 1623-27 Utica Ave., Brooklyn, NY

Maranatha SDA Church, 899 Winthrop St., Brooklyn, NY

Mount Paran Baptist Church, 12 Schafer St., Brooklyn, NY

Neighborhood Hunger Network Fall Festival, 9040 160th St., Jamaica, NY

New Life Tabernacle Church, 1476 Bedford Ave., Brooklyn, NY

Queens Elmhurst Health Fair, PS 127, 9801 25th Ave., East Elmhurst, NY

Saint Francis Xavier Church, 55 West 15th St., New York, NY

Salem Missionary Baptist Church, 305 East 21st St., Brooklyn, NY

St. John’s Lutheran Church, 81 Christopher St., New York, NY

St. Paschal Baylon Church, 112-43 198th St., Jamaica, NY

Sunrise Adult Healthcare Center, 9517 Avenue J, Brooklyn, NY

Taino Towers, 2243 3rd Ave., New York, NY

The Bowery Mission Transitional Center, 45 Avenue D, New York, NY

The Third Ave. Fair, 1180 3rd Ave., New York, NY

True Zion Gospel Temple, 145-21 Liberty Ave., Jamaica, NY

Union Baptist Church, 461 Decatur St., Brooklyn, NY

Wat Buddha Thai Thavon Vanaram Temple, 76-16 46th Ave., Elmhurst, NY

Young Women’s Leadership School, 105 East 106 St., New York, NY

We wish to acknowledge the following members of the H2H Consortium:

Venkatesh Balaji

Larry Breindel

Agnes Chayka

Paul J. Christos

Daisy Donnellan

Neville Dusaj

Kenny Wu Feng

Julia Klein

Kaymisha Knights

Marvin Yu Him Lo

Jonathan Moreno

Patrick O’Connor

Suchit Patel

Lula Mae Phillips

Jorge L. Sanchez

Gena Seraita

Lior Shtayer

Brandon Valentine

We wish to acknowledge the following past and present H2H Volunteer Medical Directors for donating so many of their weekends to this program:

William Borden, MD

Keith LaScalea, MD

Christopher Schultz, MD

Brett Ehrmann, MD

## Footnotes

1 Over the years, H2H has also been used for other innovative research studies. For example, two MD/PhD students designed a 3D-printed iPhone adapter for retinal imaging five years into the program. They secured a CTSC pilot grant to develop this device and introduce ophthalmology screenings to the H2H program. This work was published in 2022(18) and ophthalmology screenings continue to this day. In 2016, the H2H Medical Director learned that the leadership of NewYork-Presbyterian Hospital’s Dietetic Internship program sought an opportunity to provide their interns with community health experience, leading to the addition of a nutrition counseling component. At many events, participants receive culturally and medically tailored nutrition handouts, including modified recipes for cultural staples, as well as nutrition consultations from dietetic interns overseen by their licensed instructors.

2 Since 2016, with permission from the community sites, various CTSC-supported studies have attended events to recruit participants for a wide variety of research studies on topics ranging from Alzheimer’s disease to validation of new point-of-care testing devices and recruitment for the NIH AllOfUs Research Program.

3 Comprehensive screenings are held in spaces provided by community centers and faith-based organizations where participant familiarity and trust are firmly established, thereby decreasing the barriers (real and perceived) to engaging with the traditional medical establishment.

4 Program leadership has reached out to several foundations over the years and has learned that while it is not uncommon for foundations to provide community engagement programs with start-up funding to support the first year or two, identifying foundation support for ongoing, long-term funding is challenging.

5 While medical students have focused on recruiting student volunteers, the CTSC has handled IRB renewals and other administrative requirements, including: obtaining informed consent, planning events, interacting with community partners, and handling logistics (transportation, supply ordering, and funding).

6 It has not been uncommon to receive requests from students who missed annual recruitment events but learned about the opportunity from other students who attended and wanted to volunteer.

7 Cholestech LDX System

8 HgbA1c; either Bayer A1CNOW+ or Siemens DCA Vantage 2000

